# Wavelet transform analysis reveals differences between patients with impaired left ventricular systolic function and healthy individuals

**DOI:** 10.1101/2023.09.25.23296125

**Authors:** Marcin Gruszecki, Damian Kaufmann, Michał Świątczak, Krzysztof Młodziński, J. Patrick Neary, Jyotpal Singh, Jacek Rumiński, Ludmiła Daniłowicz-Szymanowicz

## Abstract

**Background:** Despite continuous progress in medical treatment, heart failure (HF) is the leading cause of hospitalizations with a high all-cause mortality in patients. Patients with a left ventricular ejection fraction (LVEF) below 50% are characterized by the highest risk of cardiovascular complications. The objective of this study was to examine how LVEF below 50% and aging impact cardiovascular physiology.

**Methods:** Sixteen males with physician diagnosed coronary artery disease and LVEF = 42 ± 6% (age 62 ± 6 years, BMI 29.1 ± 3.8kg/m^2^) and 10 healthy controls (9 male and 1 female, age 28.5 ± 9.1 years, BMI = 24.1 ± 1.2kg/m^2^) were recruited in our study. Finger photoplethysmography for blood pressure (BP) and electrocardiogram (ECG) were recorded while participants rested in a supine position. Wavelet transformations were used to analyze the amplitudes, phase coherence and phase difference of BP and ECG. The frequency intervals were separated as follows: I (0.6-2Hz), II (0.145– 0.6Hz), III (0.052–0.145Hz), and IV (0.021–0.052Hz).

**Results:** HF patients showed a decrease (p<0.05) in BP wavelet amplitude intervals III and IV in comparison to controls, and interval I for ECG. A decrease in phase coherence (p<0.01) at interval I is also found in HF patients compared to controls.

**Conclusions:** A decrease in smooth muscle cell activity and smooth muscle autonomic innervation (intervals III and IV) contributions to BP, along with a decrease in cardiac activity as shown by the wavelet amplitude in ECG, suggests altered BP and ECG function in aging HF patients. Furthermore, a decrease in the cardiac interval represents an impairment in the BP and ECG relationship in HF patients. The wavelet transform has the potential to expand our understanding of LVEF and improve diagnostic procedures and patient prognosis.

## Introduction

Heart failure (HF) is an expanding global pandemic, affecting more than 64 million people worldwide (Spencer et al. 2018). This represents a pressing public health concern due to its increasing incidence, prevalence, morbidity, mortality, and economic burden (Savarese et al. 2022). Future forecasts are even more disturbing, as HF prevalence is expected to rise in the elderly and in people living in low-to middle socio-demographic index regions (Lippi & Sanchis-Gomar 2020). Despite continuous progress in both pharmacological therapy and invasive methods (McDonagh et al. 2021) (McDonagh et al. 2021; Ahmed et al. 2021; Hussein et al. 2019), all-cause mortality in HF patients remains high with substantial economic burden on healthcare systems (Spencer et al. 2018; Savarese et al. 2022). HF is the leading cause of hospitalizations, resulting in more than 1 million admissions in both the United States and Europe (Blecker et al. 2013; Dobrowolska et al. 2021).

It is widely agreed that left ventricular ejection fraction (LVEF) is one of the most important factor that impacts the treatment and prognosis of patients with HF (Breathett et al. 2016; Park et al. 2018; Kaufmann et al. 2019). Currently, HF is categorized based on LVEF as HF with reduced (HFrEF), mildly reduced (HFmrEF), and preserved LVEF (HFpEF), according to the LVEF ranges ≤ 40%, 41–49%, and ≥ 50%, respectively. Patients with LVEF below 50% (HFmrEF and HFrEF) have similar clinical characteristics, outcomes and prognosis (Lam et al. 2014).

Decreased LVEF, marked by a reduction in the left ventricle’s ability to contract and effectively pump blood, has the potential to alter cardiac physiology. Research on the mechanistic implications of the physiological changes is limited. However, cardiovascular signals such as the arterial pressure wave and electrocardiogram are believed to have a deterministic origin, which arises from a complex interplay between physiological oscillations happening at various time scales. Analytical techniques on these cardiovascular signals have been limited as the majority have focused on analysis of the autonomic tone, being heart rate variability and baroreflex sensitivity (Daniłowicz-Szymanowicz et al. 2018 - De Ferrari et al. 2007).

The wavelet transformation analysis has emerged as a potent tool for analyzing cardiovascular signals and identifying subtle changes which may not be verified by traditional analytical methods (Stefanovska et al. 1999; Stefanovska 2007). Using the wavelet transform, it is possible to extract the deterministic aspect of the cardiovascular signals by assessing the nonlinear and time-dependent dynamics of the parameters of cardiovascular regulation, specifically, the phase coherence or difference between oscillatory components. This is a promising method, as suggested by several studies, including Smelyanskiy et al. (2005), Sheppard et al. (2012), Clemson et al. (2016) and Gruszecki et al. (2018). Such knowledge will help contribute to diagnosis and prognosis of heart failure in the future.

The purpose of this study was to use wavelet transformation technique to conduct a thorough examination of oscillatory interactions. This approach allows us to explore the impact of aging and decreased left ventricular systolic function on cardiovascular dynamics, both in terms of the central (measurement of the ECG signal) and the peripheral (measurement of the BP signal) level. Similar to the depressed systolic activity in HF patients, we hypothesize that HF patients will also exhibit depressed ECG and BP wavelet amplitudes, phase coherence and phase difference.

## Results

Table 1 shows the results of the nonparametric Wilcoxon rank sum test, which indicated a significant difference in diastolic blood pressure (DBP), systolic blood pressure (SBP), and mean arterial pressure (MAP) between the two groups. However, there was no statistically significant difference observed in HR.

**Table 1:**
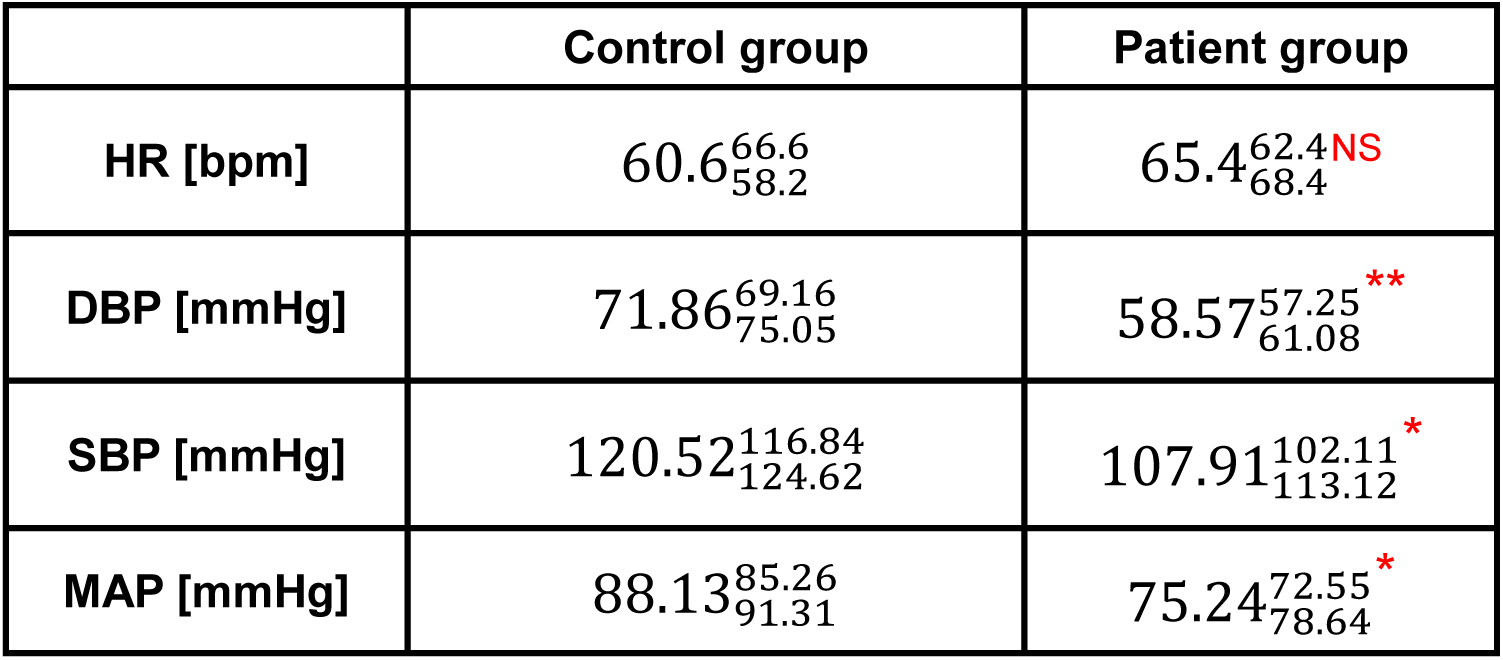
Values shown are 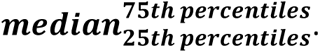. p values were estimated between the control and patient groups. HR heart rate; DBP diastolic blood pressure; SBP systolic blood pressure; MAP mean arterial pressure. *p < 0.05; **p < 0.01; ***p < 0.001.

|  | Control group | Patient group |
| --- | --- | --- |
| HR [bpm] | 60.6<sup>66.6</sup><sub>58.2</sub> | 65.4<sup>62.4</sup><sub>68.4</sub> NS |
| DBP [mmHg] | 71.86<sup>69.16</sup><sub>75.05</sub> | 58.57<sup>57.25</sup><sub>61.08</sub> ** |
| SBP [mmHg] | 120.52<sup>116.84</sup><sub>124.62</sub> | 107.91<sup>102.11</sup><sub>113.12</sub> * |
| MAP [mmHg] | 88.13<sup>85.26</sup><sub>91.31</sub> | 75.24<sup>72.55</sup><sub>78.64</sub> * |

Figure 1 illustrates the results of the wavelet transform (WT) amplitude for a single representative volunteer from the control group (panels a, b) and the patient group (panels c, d). The wavelet transform was applied to both BP (panels a, c) and ECG (b, d) signals. The entire duration of the collected signals reveals prominent cardiac oscillations at approximately 1 Hz. Additionally, there are observable oscillations at lower frequencies for both groups.

**Figure 1.**
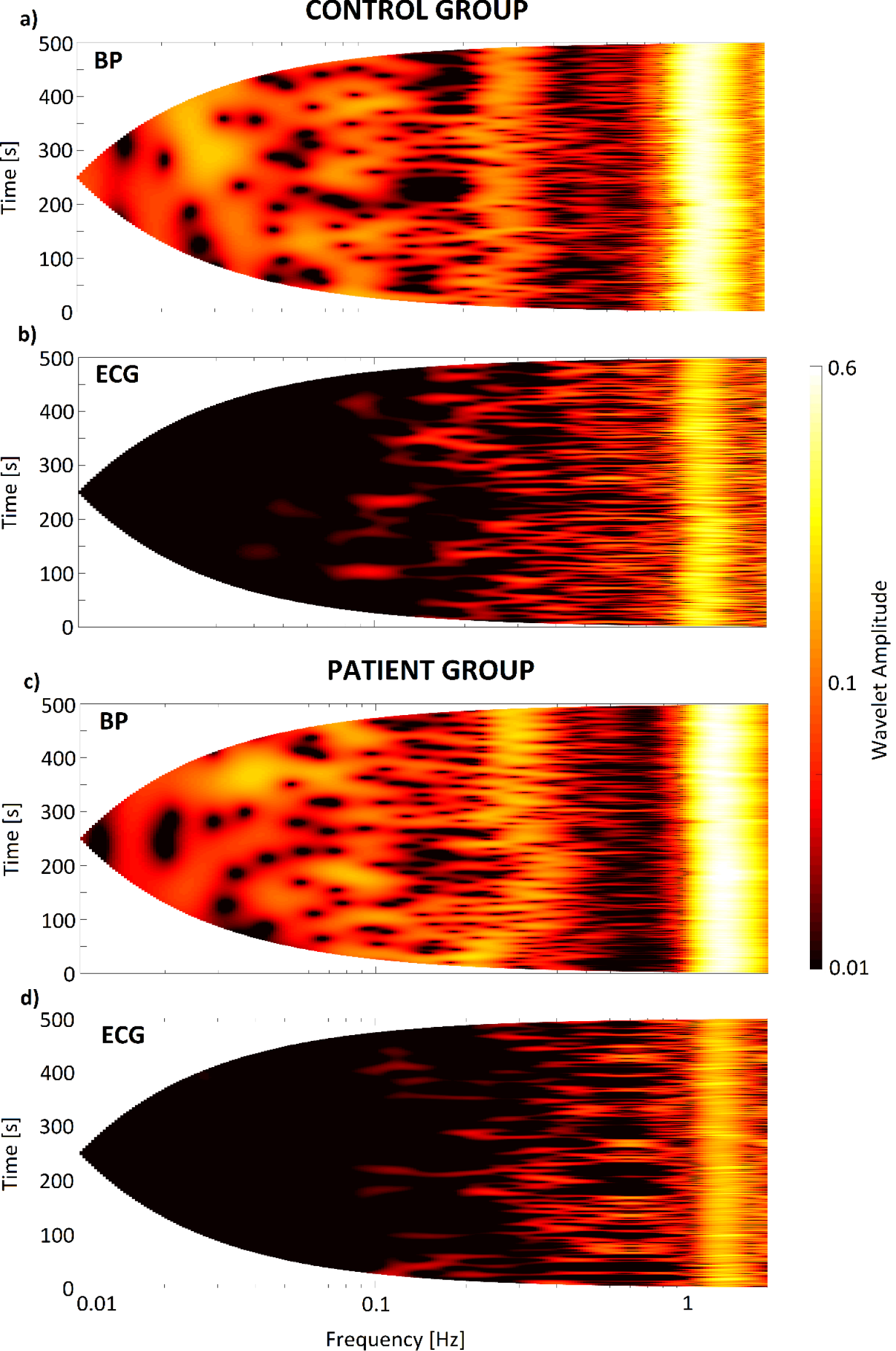
*Wavelet amplitude of recorded signals: BP (a, c) and ECG (b, d) are shown for one representative volunteer. The top two panels (a and b) illustrate the results for one of subject in the control group, while the bottom two panels (c and d) show the results for one of subject in the patient group*.

Figure 2 represented the comparison of frequency content between the control and patient groups using the median of time-averaged amplitude of wavelet transforms. Our analysis was augmented by incorporating four distinct frequency intervals, previously described by Stefanovska et al. (1999) and Gruszecki et al. (2018), which correspond to various physiological functions. Intervals I and II (0.6–2 Hz and 0.145– 0.6 Hz, respectively) relate to cardiac and respiratory function, while interval III (0.052– 0.145 Hz) usually pertains to smooth muscle cell activity and interval IV (0.021– 0.052 Hz) has been proposed to reflect smooth muscle autonomic innervation (Stefanovska et al. 1999). Additionally, we calculated the p value to identify significant differences between the results of the control and patient groups. We observed statistically significant differences between the two groups for III and IV intervals in BP signal, and for the I frequency interval in ECG signal.

**Figure 2:**
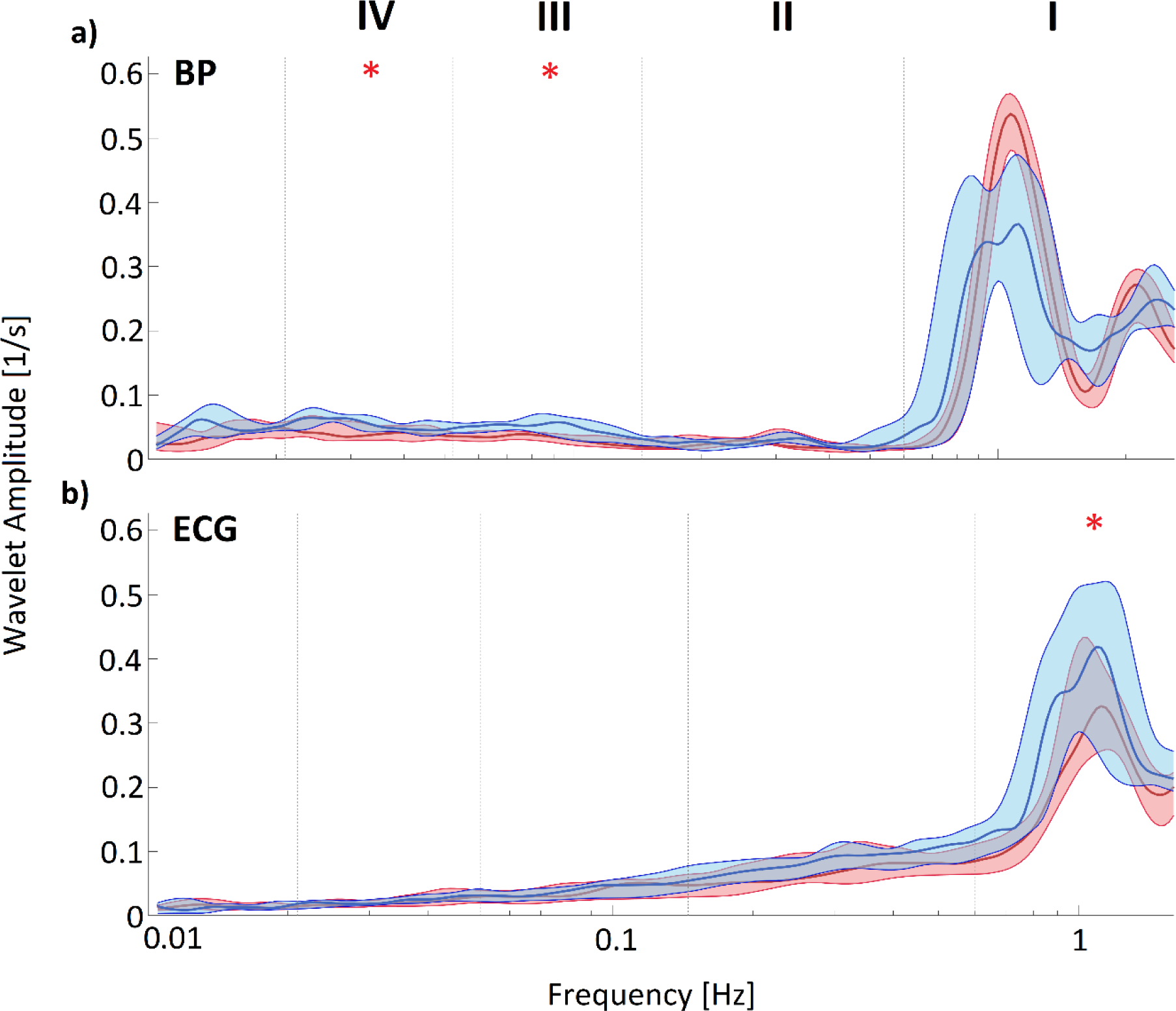
*Shown in panels a) and b) thick lines are the median time-averaged wavelet transforms of signals recorded from subjects in the patient and subjects in the control groups, respectively. Thick lines in red and blue represent the patient and control groups, respectively, while the shaded areas indicate the inter-quartile range (25th to 75th percentiles). Significant differences between groups are indicated by asterisks: *p < 0.05; **p < 0.01; ***p < 0.001. Panel a) shows the results for the BP signal, while panel b) displays the results for the ECG signal*.

The phase coherence value was deemed to be significant when it exceeded the 95th percentile of 190 inter-subject surrogates (generated from 2-permutations of 20 subjects). Statistically significant difference (p<0.01) was found in phase coherence at the cardiac interval (Figure 3a). No statistically significant differences were found in phase differences (Figure 3b).

**Figure 3:**
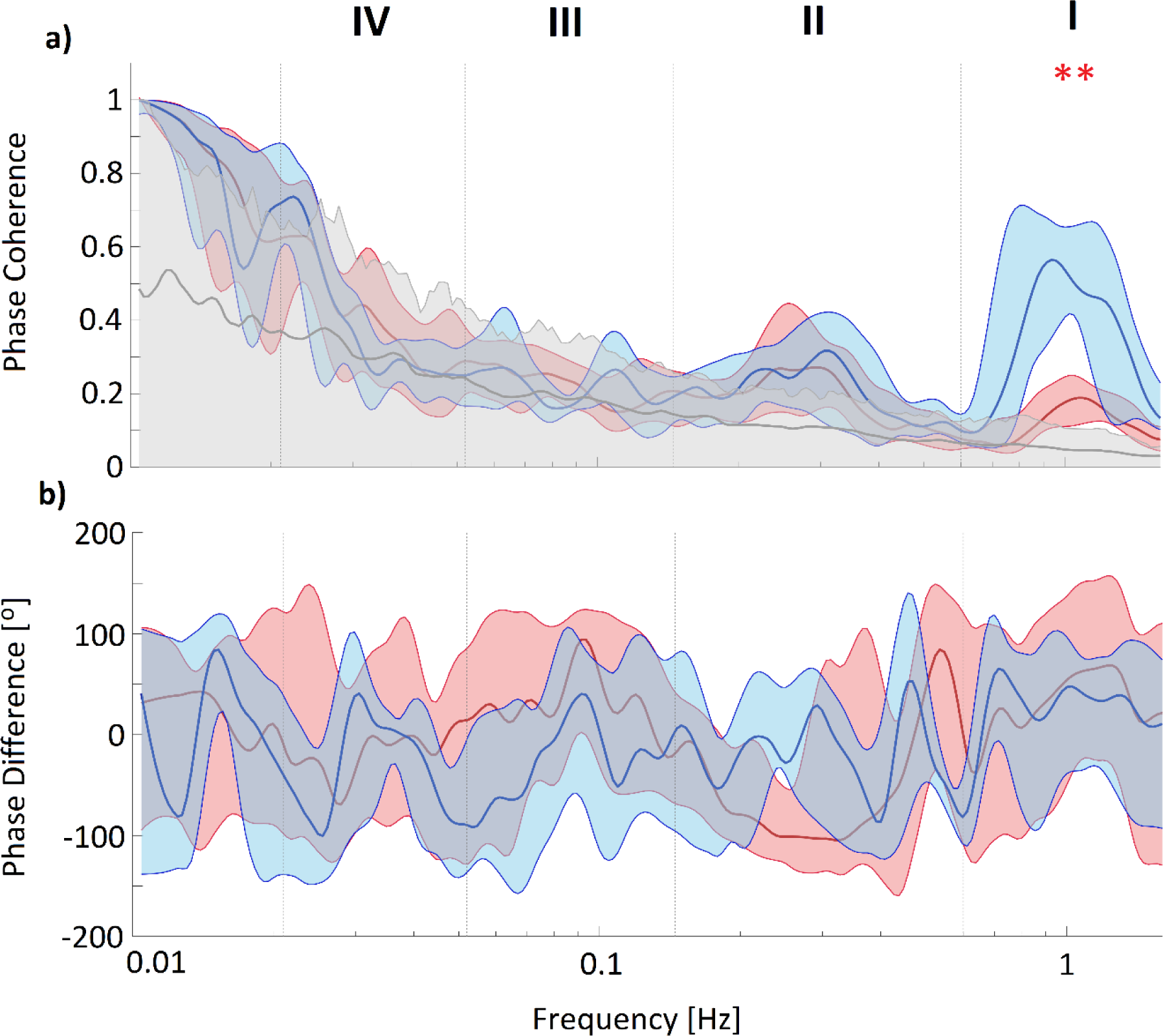
*Panel a) displays the median wavelet phase coherence between BP and ECG, while panel b) shows the median phase difference between the same two signals. The thick coloured lines indicate the median values, and the coloured shading represents the interquartile range (25th to 75th percentiles). Coherence below the 95th percentile of the surrogates is not considered significant and is shown by the light grey line and shading. The red and blue lines and shaded areas correspond to the patient and control groups, respectively. Significant differences between groups are indicated by asterisks: *p < 0.05; **p < 0.01; ***p < 0.001*.

In addition, we estimated the time evolution of heart rate (HR) by fitting a line in the middle of the cardiac mode (represented by the dark red line in panel a of Figure 4).

**Figure 4.**
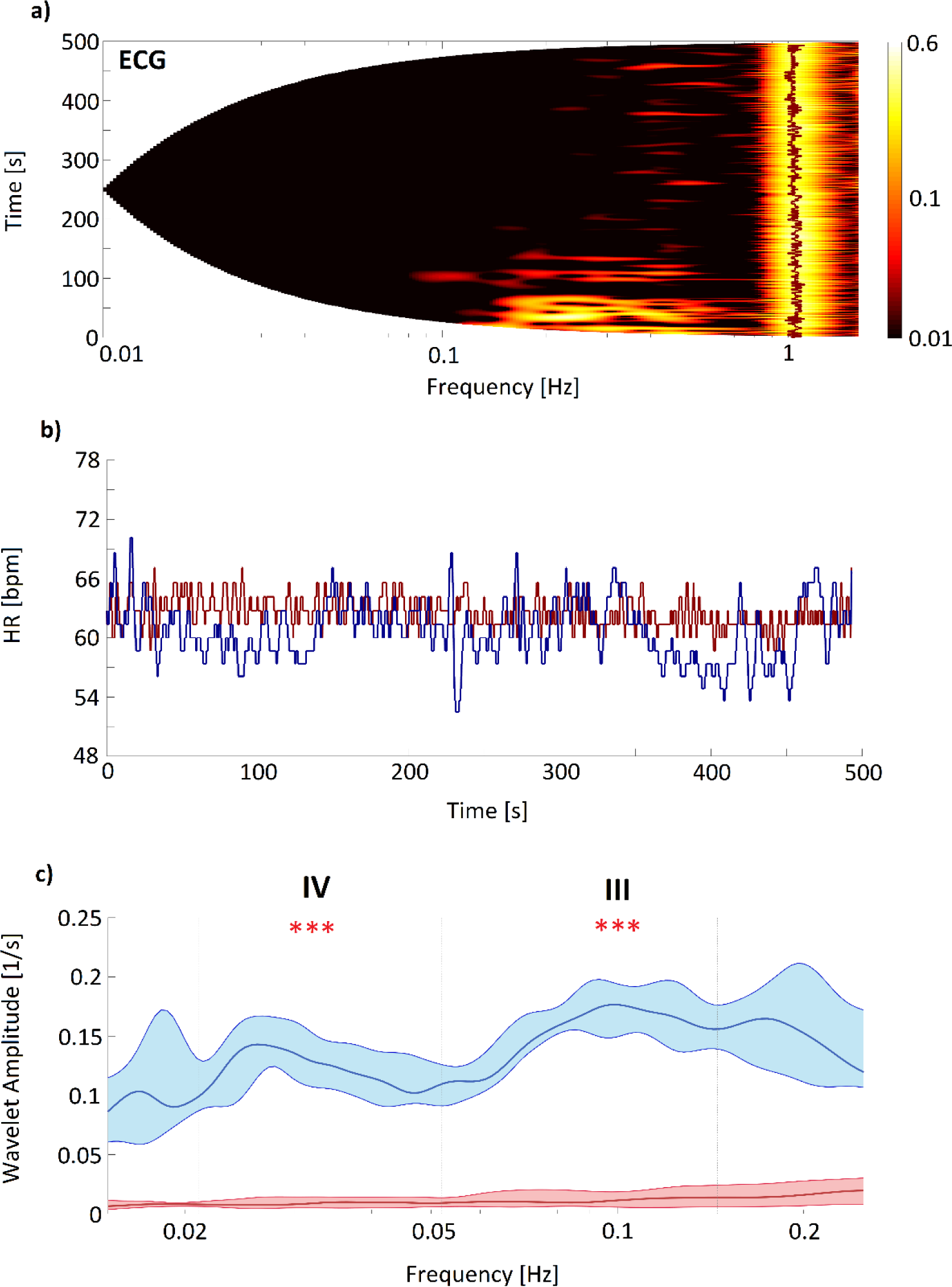
*Panel a) displays the wavelet transform results for the ECG signal collected from one of the volunteers in the patient group. The red line illustrates the best fit to a cardiac mode. Panel b) shows the time evolution of the frequency of the cardiac mode, with the red (blue) line corresponding to the patient (control) group. In panel c), the thick lines represent the median time-averaged wavelet transform, with the shaded areas indicating the interquartile range (25th to 75th percentiles). The colours red and blue correspond to the patient and control groups, respectively. Significant differences between groups are indicated by asterisks: *p < 0.05; **p < 0.01; ***p < 0.001*.

Figure 4, panel b, displays the HR time evolution for two subjects, one from the patient group (represented by the dark red line) and the other from the control group (represented by the dark blue line). Additionally, we calculated the median of time-averaged amplitude of wavelet transforms for the HR signal in both groups, which is shown in panel c of Figure 4. Our analysis indicated that only two frequency intervals (III and IV) showed statistically significant differences between the control and patient groups.

## Discussion

Under resting conditions, we have been able to investigate changes in the dynamic cardiovascular processes associated with aging and reduced LVEF by analyzing the wavelet amplitude, phase coherence and phase difference. Wavelet transformation is a very useful technique for analyzing physiological signals that contain complex, non-stationary oscillations, such as heart rate variability and blood pressure fluctuations.

The presence of 0.1 Hz oscillation in vessel radius has been widely documented in humans. The application of wavelet transform has allowed the detection of these oscillations in BP signals (Gruszecki et al. 2018; Gruszecka et al. 2021), as well as signals obtained from laser Doppler flowmetry measurements (Kvernmo et al. 1998; Kvandal et al. 2003; Stefanovska et al. 2007). Despite extensive discussions in the literature, there is still no consensus regarding the origin of these oscillations. Some researchers attribute these oscillations to the sympathetic nervous system (Stauss et al. 1998; Cevese et al. 2001), linking them to baroreceptor activity that regulates heart rate, thus controlling and stabilizing blood pressure. Conversely, other studies suggest that the 0.1 Hz oscillation arises directly from spontaneous contractions of pressure-sensitive pacemaker cells within the smooth muscle of the arterial walls (Johnson 1991, Söderström et al. 2003), independent of the sympathetic system. Previous studies on skin-flaps, conducted under either local or general anaesthesia, have shed further light on the origin of oscillations observed in the vascular myocytes. These studies, such as those by Söderström et al. (2003) and Landsverk et al. (2006, 2007), have shown that myogenic oscillations (at around 0.1 Hz) can be distinguished from slower sympathetic oscillations (at around 0.04 Hz) when sympathetic nerves are either absent or temporarily blocked. The myogenic oscillations are believed to be spontaneously activated by ionic conductance in the smooth muscle cells or stimulated by sympathetic inflow, and they contribute to the regulation of vascular stiffness. These findings may explain the impaired myogenic activity (interval III) observed in the patient group for both the BP signal (Fig. 2 panel a) and HR signal (Fig. 4 panel c), i.e., potentially due to an increase in vessel stiffness among older people (Levy et al. 2001; Ticcinelli et al. 2017).

Blood pressure, blood flow, and heart rate variability signals exhibit a periodicity around 0.03 Hz, which is attributed to sympathetic nerve activity. This was confirmed by Stefanovska et al. (1999) using wavelet analysis. In humans, microvascular flaps lacking sympathetic innervation exhibit reduced spectral amplitude in the low-frequency range of 0.021-0.051 Hz compared to adjacent skin, as shown by Söderström et al. (2003) and Landsverk et al. (2006, 2007). After brachial plexus block and during general anesthesia, Landsverk et al. (2006, 2007) observed a significant reduction in spectral amplitude of laser Doppler flowmetry (LDF) signals in the lower forearm skin compared to the control group. We observed a significant decrease in the amplitude of wavelet transforms in the patient group compared to the control group (Figure 2a and Figure 4c), which is consistent with earlier findings by Bernjak et al. (2008). They showed a reduction in the absolute spectral amplitude of LDF signals in patients with congestive HF compared to healthy controls. This reflects impaired smooth muscle autonomic innervation for patients with impaired left ventricular systolic function.

The significant reduction in phase coherence between BP and ECG signal in the cardiac frequency interval (I; 0.6-2Hz) for the patient group could be explained by the impaired left ventricular systolic function of the heart, specifically the reduced LVEF. This leads to phase changes between blood pressure and ECG that affect the coherence of the two signals at the cardiac frequency interval. For healthy participants, the phase coherence between BP and ECG is not disturbed and remains intact.

The wide variety of drugs used to treat participants from the patient group included in our study presents a challenge for researching treated patients. As noted in the Subject subsection, this diversity of drugs makes it difficult to determine which drugs are more or less effective. This limitation of our study prevents us from drawing definitive conclusions on this matter. However, despite this limitation, our study demonstrates that even in a group of patients with optimally treated reduced LVEF, there are still impairments in cardiac, myogenic, and neurogenic functions. The same methodology used in our study can be applied to evaluate the effects of individual drugs or to conduct longer-term follow-up studies on LVEF treatment. This approach could contribute to the development of new medications.

## Conclusions

In conclusion, by examining the deterministic characteristics of BP and ECG signals and analyzing time-dependent parameters, we have obtained clinically relevant findings pertaining to aging and the decline of left ventricular systolic function. Unique to our study is the exploration of phase coherence between BP and ECG. The observed decrease in coherence within the patient group’s cardiac frequency interval provide new clinical marker for patient with decreased LVSF. Medical treatment is evidently unable to correct this defect. There is a need to introduce a new treatment guidelines and medications to increase value of this parameter for patient with reduced LVEF.

## Materials and Methods

### Subjects

Prior to the experiment, participants were instructed to abstain from alcohol for at least 24 hours, and from tea, coffee, nicotine, cocoa, and any food and beverages containing methylxanthine for at least 12 hours. Intense exercise training was prohibited for at least 6 hours prior to testing, and all participants were asked to empty their bladder within 30 minutes of testing (Ellingson et al., 2022). Before each procedure, participants rested for 10 minutes in a comfortable chair in a quiet room.

#### Patient group

The patient group consisting of 16 patients with stable ischemic HF (documented by prior myocardial infarction, percutaneous coronary intervention, or coronary artery by-pass grafting) and LVEF ≤ 50%. Additional inclusion criteria included sinus rhythm, optimal pharmacological therapy, stable clinical condition for at least 3 months before enrollment, and without significant features of hypervolemia at the moment of enrollment. The patients were excluded if they were younger than 18 years old, had a history of sustained ventricular arrhythmia (ventricular tachycardia or ventricular fibrillation) or cardiac arrest, were in New York Heart Association functional class IV, permanent atrial fibrillation or flutter, permanent second- or third-degree atrioventricular block, had an implanted pacemaker, had clinical features of coronary instability at the moment of enrolment, a revascularization (coronary angioplasty or/and surgery by-pass) within 3 months prior to the study, or incomplete coronary revascularization status (scheduled control coronarography, coronary angioplasty or surgery by-pass), clinical evidence of autonomic neuropathy, concomitant terminal disease and non-cardiologic comorbidities with a potentially unfavorable effect on survival. Baseline characteristics of the patients group were as follow: 15 out of 16 patients were men with a mean age of 62 ± 6 years and BMI of 29.1 ± 3.8 kg/m^2^. Most of the patients underwent myocardial infarction (88%), had hypertension (75%), hypercholesterolemia (69%) and were active smokers (75%). Diabetes and atrial paroxysmal fibrillation were present in 25%. Regarding pharmacotherapy 94% of patients received beta-blockers, 88% had angiotensin-converting enzyme inhibitors/angiotensin receptor blockers, 94% patients were treated with statins, 88% with antiplatelet drugs, 50% with mineralocorticoid receptor blockers and 25% with diuretics and anticoagulants. The study was conducted in accordance with the Helsinki recommendations and was approved by the Ethics Committee of Medical University of Gdansk (NKBBN/864/2022-2023).

#### Control group

Ten control participants (9 males and 1 female, age 28.5 ± 9.1 years, BMI = 24.1 ± 1.2 kg/m^2^), who were healthy, non-smokers, and over 18 years of age, participated in the study. The Ethics Committee of University of Regina (REB#2017-013) approved this study and the experimental protocol. All control participants provided signed informed consent forms.

#### Experimental design

For both groups (control and patient) testing was carried out between 08.00 a.m. and 1.00 p.m. in a quiet, comfortable room set to a temperature of 18-20°C with minimal ambient lighting. Once all the necessary medical research equipment was attached to the volunteer participant (see “Measurements” section below), they were directed to lie down on a bed with a headrest for 15 minutes of the stabilization period (provided with a blanket if necessary).

#### Measurements

During the experiment the researchers used a Finometer (Finapres 2300, Ohmeda) to measure the participants’ blood pressure (BP). The Finometer employs a finger-cuff to monitor beat-to-beat blood pressure from the left middle finger. The heart rate (HR) was determined from an electrocardiogram (ECG) signal, with the ECG ground electrode placed on the left anterior superior iliac spine and the two primary leads located under the middle portion of each clavicle (Gruszecka et al. 2020).

The BP and ECG signals were obtained at a sampling rate of 300 Hz and then processed for analysis. Pre-processing steps involved detrending and normalization of the signals by subtracting their mean and dividing by their standard deviation (Gruszecki et al. 2018). Additionally, the signals were downsampled to a lower frequency of 20 Hz to reduce the amount of data while retaining important biological waveform features of the ECG and BP signals.

#### Wavelet transform

Wavelet analysis was employed to identify and analyze the physiological mechanisms underlying the oscillations in the cardiovascular system. The wavelet transform is a powerful mathematical tool used in signal processing that enables the transformation of a signal from the time domain to the time-frequency domain. Unlike traditional Fourier analysis, which provides information about the frequency content of a signal but not its temporal evolution, wavelet analysis allows us to examine how the frequency content of a signal changes over time. The definition of the wavelet transform is:

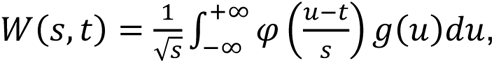

where *W*(*s, t*) is the wavelet coefficient, *g*(*u*) is the time series and φ is the Morlet mother wavelet, scaled by factor *s* and translated in time by *t*. The Morlet mother wavelet is defined by the equation:

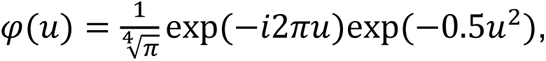

where 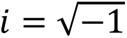. The Morlet wavelet is a commonly used wavelet in signal processing due to its Gaussian shape, which allows for good localization of events in time and frequency domains (Bernjak et al. 2012). This means that the Morlet wavelet can accurately capture changes in frequency content over time, making it a valuable tool for analyzing complex signals with non-stationary oscillations, such as those seen in physiological systems. The Gaussian shape of the Morlet wavelet also helps to reduce spectral leakage, which can occur when analyzing signals with sharp, sudden changes in frequency content. When the Morlet wavelet is used for wavelet analysis, the resulting wavelet coefficients are complex numbers in the time-frequency plane:

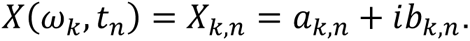

They define the instantaneous relative phase,

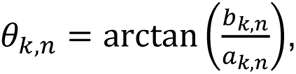

and the absolute amplitude,

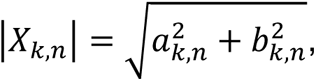

for each frequency and time.

During the measurement, heart may create phase modulations. A mathematical tool to find the relationship between the phases of two signals is the wavelet phase coherence (WPCO). WPCO enables us to determine whether the oscillations detected are significantly correlated over time and can be a valuable tool for investigating the complex dynamics of heart signals and their relationship to other physiological signals. To estimate the WPCO we used the following expression (Lachaux et al. 2002):

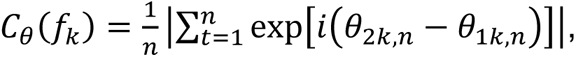

where 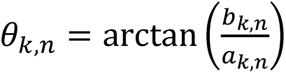 is an instantaneous measure of phases at each time t_n_ and frequency *f_k_* for both signals. WPCO approaching zero when two oscillations are unrelated and their phase difference continuously changes with time. When two oscillations are related, their phase difference remains constant with time, and the wavelet phase coherence (WPCO) approaches 1.

Additionally, we can obtain information about the phase lag of one oscillator relative to the other by calculating the phase difference Δθ_*k*_ defined as:

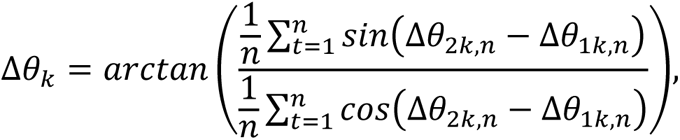

where Δθ_*k*_ ∈ (−180°, 180°).

#### Statistical analysis

Nonparametric statistical tests were used to avoid assumptions of normality. Specifically, the Wilcoxon rank sum test was used to assess the significance of differences in median values between the control and patient groups. Significance was set at p<0.05.

To determine the statistical significance of the estimated values of phase coherence, we employed the surrogate data testing method (Lancaster et al. 2018). Since fewer cycles of oscillations naturally occur at lower frequencies, there can be artificially increased wavelet phase coherence, even in cases where there is none. The surrogate analysis enables us to identify a significance level above which the phase coherence can be deemed physically meaningful. For this purpose, we used intersubject surrogates (Sun et al. 2012), which assume that the signals collected from different subjects are independent but have similar characteristics. The threshold value of phase coherence obtained at each frequency was then compared to the surrogate threshold, and coherence values above the threshold were considered statistically significant.

## Funding

This work has been partially supported by subsidy Funds of Electronics, Telecommunications and Informatics Faculty, Gdansk University of Technology.

## Data Availability

The data presented in this study are available on request from the corresponding author. The data are not publicly available due to privacy.

## Acknowledgements

We would like to thank all of the volunteer participants that contributed their valuable time to these experiments.

## Competing interests

The authors declare no competing interests.

## Author Contributions

Conceptualization, L.D.S. D.K. and M.G.; methodology, M.G.; software M.G.; validation, M.G.; formal analysis, M.G.; investigation, M.G. resources, L.D.S., D.K., M.Ś., K.M., J.P.N., J.S., J.R.; data curation, M.G.; writing—original draft preparation, M.G., L.D.S. and D.K.; writing—review and editing and visualisation, M.G., L.D.S., D.K., M.Ś., K.M., J.P.N., J.S., J.R.; and funding acquisition, J.R.

All authors have read and agreed to the published version of the manuscript.

## Notes

### Competing Interest Statement

The authors have declared no competing interest.

### Author Declarations

The study was conducted in accordance with the Helsinki recommendations and was approved by the Ethics Committee of Medical University of Gdansk (NKBBN/864/2022-2023). The Ethics Committee of University of Regina (REB#2017-013) approved this study and the experimental protocol.

